# Who is most at risk of dying if infected with SARS-CoV-2? A mortality risk factor analysis using machine learning of COVID-19 patients over time in a large Mexican population

**DOI:** 10.1101/2023.01.17.23284684

**Authors:** Lauren D. Liao, Alan E. Hubbard, Juan Pablo Gutiérrez, Arturo Juárez-Flores, Kendall Kikkawa, Ronit Gupta, Yana Yarmolich, Iván de Jesús Ascencio-Montiel, Stefano M. Bertozzi

**Affiliations:** University of California, Berkeley, School of Public Health, Berkeley, CA, USA; Center for Policy, Population & Health Research, School of Medicine, Universidad Nacional Autónoma de México, Mexico City, Mexico; Micron Technology, Boise, ID, USA; Instituto Mexicano del Seguro Social, CDMX, México; University of Washington School of Public Health, Seattle, WA, USA; Instituto Nacional de Salud Pública, Cuernavaca, MOR, México

**Keywords:** mortality, death, COVID-19, biostatistics, international health

## Abstract

**Background:** COVID-19 would kill fewer people if health programs can predict who is at higher risk of mortality because resources can be targeted to protect those people from infection. We predict mortality in a very large population in Mexico with machine learning using demographic variables and pre-existing conditions.

**Methods:** We conducted a population-based cohort study with over 1.4 million laboratory-confirmed COVID-19 patients using the Mexican social security database. Analysis is performed on data from March 2020 to November 2021 and over three phases: (1) from March to October in 2020, (2) from November 2020 to March 2021, and (3) from April to November 2021. We predict mortality using an ensemble machine learning method, *super learner*, and independently estimate the adjusted mortality relative risk of each pre-existing condition using targeted maximum likelihood estimation.

**Results:** Super learner fit has a high predictive performance (C-statistic: 0.907), where age is the most predictive factor for mortality. After adjusting for demographic factors, renal disease, hypertension, diabetes, and obesity are the most impactful pre-existing conditions. Phase analysis shows that the adjusted mortality risk decreased over time while relative risk increased for each pre-existing condition.

**Conclusions:** While age is the most important predictor of mortality, younger individuals with hypertension, diabetes and obesity are at comparable mortality risk as individuals who are 20 years older without any of the three conditions. Our model can be continuously updated to identify individuals who should most be protected against infection as the pandemic evolves.

**Key messages:** *What is already known on this topic:* Studies for Mexico and other countries have suggested that pre-existing conditions such as renal disease, diabetes, hypertension, and obesity are strongly associated with COVID-19 mortality. While age and the presence of pre-existing conditions have been shown to predict mortality, other studies have typically used less powerful statistical approaches, have had smaller sample sizes, and have not been able to describe changes over time.

*What this study adds:* This study examines mortality risk in a very large population (> 60 M); it uses powerful ensemble machine learning methods that outperform regression analyses; and it demonstrates marked changes over time in the degree to which different risk factors predict mortality.

*How this study might affect research, practice or policy:* Because we show an important improvement in predictive performance over traditional regression analyses, and the ability to update estimates as the pandemic evolves, we argue that these methods should be much more widely used to inform national programming in Mexico and elsewhere. Programs that assume that predictive models don’t change over time as variants emerge and as pre-existing immunity evolves due to vaccination and prior infection will not accurately predict mortality risk.

## Introduction

The probability that someone infected with SARS-CoV-2 dies has varied enormously over time, among countries, and among population groups within countries. Interest in understanding who is at a higher risk of death has grown as this heterogeneity became more apparent. Identifying people at higher risk of severe disease and death will help health systems better respond and focus prevention resources on protecting them. We examine Mexico, a country with a very high reported case-fatality rate (4.7%) among those who have laboratory-confirmed coronavirus disease 2019 (COVID-19) as of September 23, 2022 [1].

Previous analyses in Mexico have found diabetes, obesity, hypertension, immunosuppression, and renal disease to be significant risk factors along with age and sex. Multiple authors have identified obesity and diabetes as important risk factors for mortality [2–5]. Escobedo de-la Peña et al. also found a strong association with hypertension, which is consistent with results from Giannouchos et al. [5,6]. Late-stage chronic kidney disease, although less prevalent, has also consistently been identified as a COVID-19 mortality risk factor. Older/ male patients tend to have higher mortality risks than younger/ female patients [3,5,6]. In a previous analysis, we found interactions between those comorbidities, suggesting a synergic effect when having more than one of diabetes, hypertension, and obesity (larger odds ratio when reporting the 3 conditions vs. one or two) [7]. We also found that the odds ratio increased by age group with those over age 80 having 30-fold the risk of those 20 to 29 [7]. One important consideration is that the prevalence of diabetes and hypertension is positively associated with age, so it has not been clear how this interaction is related to mortality risk. A more adaptive analysis performed by Martínez-Martínez et al. developed a prediction model for severity of COVID-19, defined by hospitalization and/or mortality. They examined the relationship of 14 variables with hospitalization and mortality using interaction terms and splines to account for non-linear relationships [8].

The pattern of age, sex, and comorbidities being associated with higher mortality risk is not specific to Mexico, and the global literature on such associations is extensive. Researchers have identified old age, diabetes, obesity, chronic renal failure, and congestive heart failure to be strongly associated with severe infection amongst both sexes in the Spanish population [9]. Researchers in Brazil showed that older age, male, kidney disease, obesity and/ or diabetes are strong predictors of mortality amongst other comorbidities such as chronic liver disease, immunosuppression, and cardiovascular disease [10,11]. Another study used United Kingdom Biobank data and showed that pre-existing dementia, diabetes, chronic obstructive pulmonary disease (COPD), pneumonia, and depression were positively associated with risk of hospitalization and death [12]. An analysis from France found age, diabetes, hypertension, obesity, cancer, and kidney and lung transplants to be associated with risk of COVID-19-related hospitalization and mortality, among others [13]. A Canadian study reported dementia, chronic kidney disease, cardiovascular disease, diabetes, COPD, severe mental illness, organ transplant, hypertension, and cancer to be significant predictors of mortality [14]. Our goal in this study is not only to predict mortality using demographic factors and comorbidities, but to show how those predictions change over time in this rapidly evolving pandemic.

Although mortality risk estimation and risk factor identification have been examined in prior studies, we are concerned about the statistical validity and interpretation of the standard methods. A commonly used prediction tool, logistic regression, assumes a linear relationship of predictors against the log odds of mortality risk, but this logit-linear assumption will lead inevitably to biased estimates of risk (either under- or over-predict the risk) for subsets of the population. We instead used flexible, data-adaptive methods that can capture non-linearities in the dose-response, such as potential nonlinear interactions between the predictors (e.g., the potential interaction of age and diabetes on predicting death) [15,16]. The better the model fits the study population; the more likely estimates are closer to the true joint relationship of mortality and risk factors.

We included pre-existing conditions, demographic variables, the Mexican state where the patient was treated, and the month that the patient initiated care to fit our prediction algorithm. We conducted the analysis using an ensemble machine learning algorithm, super learner, to form optimal combination of predictions from multiple machine learning methods [15,16]. We also estimated the comparative importance of variables for mortality risk prediction (holding all other variables constant) by nonparametrically estimating quantities inspired by causal parameters (parameters that compare so-called counterfactual distributions, in our case, causal relative risks). The statistical goal is to estimate and provide robust inference for impact estimates of the predictors without the arbitrary modeling assumptions that characterize the great majority of prior work [17].

## Methods

### Study population and design

The study population is drawn from the Mexican Social Security Institute (IMSS), a vertically integrated insurance and health system that provides coverage for over 60 million private sector employees and their families, including their parents, children and spouse. IMSS also provided care as part of the COVID-19 response for some non-beneficiaries, who are also included in the dataset.

The data were recorded from March 1st, 2020, to November 3rd, 2021 in a platform called SINOLAVE. They reflect the entire population of 4,482,292 patients who were registered as receiving care for suspected COVID-19 at an IMSS facility. The dataset and the data entry process have been described previously [18]. The demographic variables include age, sex, insured by IMSS, and indigenous status. The data contains pre-existing conditions reported by the patient or the family at presentation: asthma, cardiovascular disease, chronic liver disease, chronic obstructive pulmonary disease, diabetes, hemolytic anemia, human immunodeficiency virus, hypertension, immunosuppression, neurological disease, obesity, cancer, renal disease and tuberculosis, as well as whether the patient currently smokes. Patients were asked at presentation about their pre-existing health conditions; these were not ascertained with reference to the patient’s medical record, even for those patients insured by the IMSS. The data also includes the Mexican state in which the patient received care, COVID-19 test results (from both polymerase chain reaction (PCR) tests and antigen tests), the month that the patient initiated care, and mortality. In addition, we extracted a different dataset from the National Council of Science and Technology to determine the dominant circulating variant in each month [19]. A short summary can be found in **Table 1 (Supplemental Table S1)**. We define COVID-19 positive as a positive PCR or antigen test.

**Table 1.**
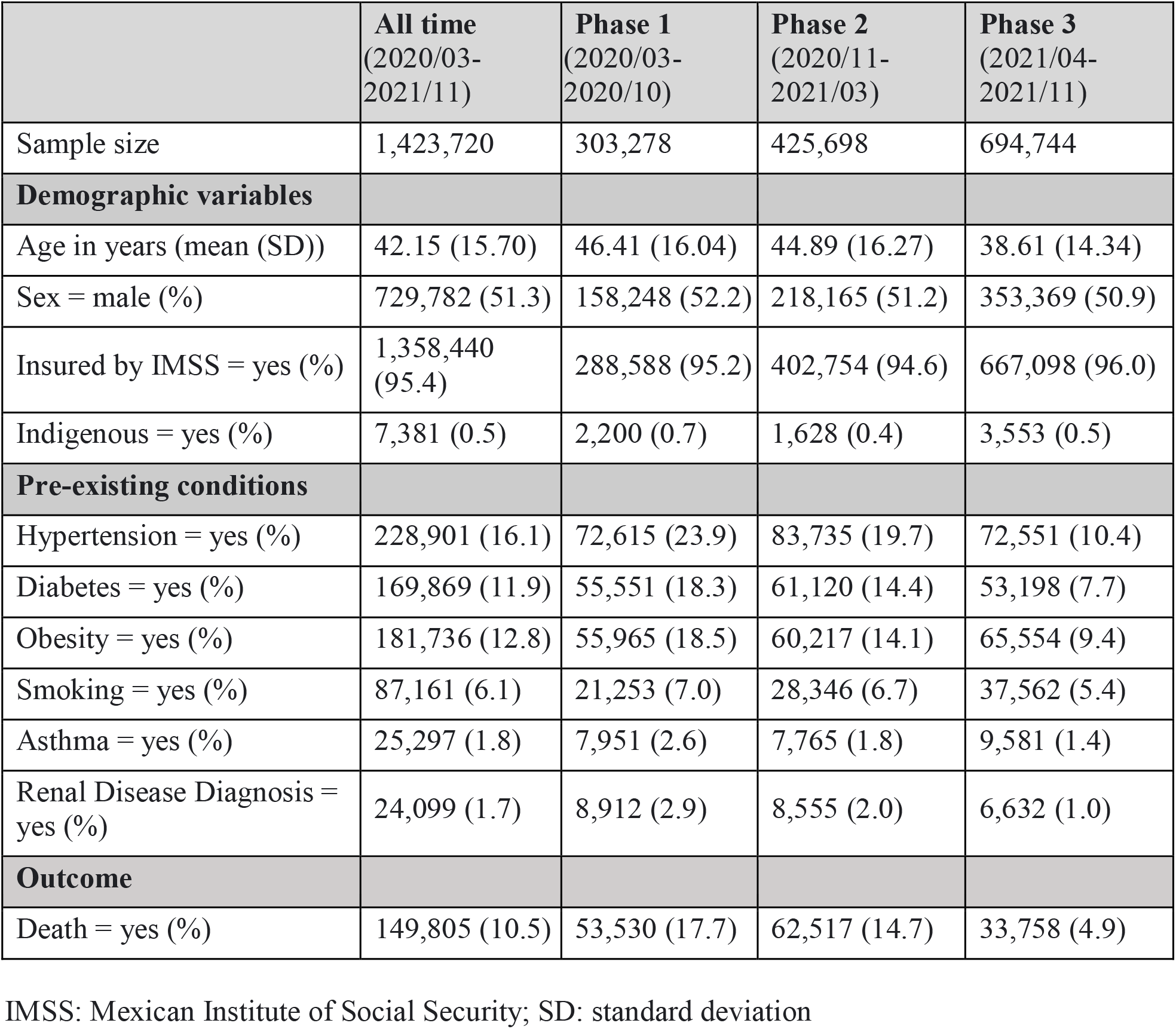
Summary table of baseline variables and pre-existing conditions.

From the full data set, we generated an analytic sample (n = 1,423,720) **(Supplemental Figure S1)**. We exclude those under the age of 20 years, those without any positive COVID-19 test result from either the PCR or antigen tests, and those with unknown pre-existing conditions. We also create a phase variable that corresponds to changes in the epidemic curve into three: phase 1 is from March 1st, 2020, to October 31st, 2020, phase 2 is from November 1st, 2020, to March 31st, 2021, and phase 3 is from April 1st, 2021, to November 3rd, 2021 as previously described [18].

### Statistical analysis

#### Mortality risk prediction using super learner (SL)

We predict mortality risks with SL [15,16], using predictors: pre-existing conditions, demographic variables, the Mexican state where the patient was treated, and the month that the patient initiated care. SL combines a set of user-supplied machine learning algorithms, which includes both simple, parametric fits and flexible algorithms, to create an optimally-weighted combination. This optimal fit is found by creating a combination of algorithms that minimize the cross-validated risk (in our case, the negative log-likelihood). SL has the property that asymptotically it will perform at least as well as the best fitting algorithm in the library [15,16]. Thus, it is important to include a diverse and large set of learners as candidates to ensure the model can fit complex patterns if warranted, but also, simpler, parametric models if simpler fits are sufficient. The following learners were included in the SL library: Bayesian additive regression trees [21], Bayesian generalized linear model [22], elastic net regression [23], empirical mean, generalized additive model [24], least absolute shrinkage and selection operator regression [25], logistic regression, multivariate adaptive regression splines [26], random forest [27], ridge regression [28], and extreme gradient boosting algorithms [29]. We estimate the prediction performance, via the AUC, and derive a 95% confidence interval for the estimated AUC [30]. We compare the SL fit using all predictors listed above to a logistic regression with only age entered as a linear term. We compute the AUC for the resulting SL/logistic regression fits with 3-fold cross validation on the 80%, both on the same data used to estimate SL/logistic regression models (training AUC), as well as a more realistic assessment by using the test set – the left-out 20% of the available data (testing AUC).

To interpret the final prediction model generated by the SL fit, we use the permutation-based variable importance measure to identify variables that influence the SL model’s prediction [27]. This is performed by permuting the predictor variables one at a time (keeping the other variables fixed) and measuring the magnitude of the decline on the predictive performance (as measured by the change in the average negative log-likelihood). This provides a list of variables ranked by the relative importance to prediction fit but does not provide information on the variable impact on mortality, which led us to another measure of relative risk (RR) using targeted maximum likelihood estimation (TMLE).

#### Pre-existing condition relative risk estimate through targeted maximum likelihood estimation

For pre-existing conditions, we estimated a different variable importance measure that is not focused on prediction accuracy but on estimating potential impacts of pre-existing conditions on mortality risk. The impact is estimated by the RR of adjusted means (adjusted for baseline confounders) for the population if everyone had the specific pre-existing condition of interest (the numerator) versus the same population where no one has the specific pre-existing condition (the denominator). To estimate RRs, we used cross-validated targeted minimum-loss-based estimation (cross-validated TMLE). TMLE is a semiparametric, substitution estimator that has shown to be asymptotically efficient (unlike the inverse probability of treatment-weighting estimators [31]). It also has some robustness advantages over other semiparametric efficient approaches, such as augmented inverse probability weighting. TMLE estimates parameters that, under certain assumptions, can be interpreted as potential causal impacts of these factors on mortality, in our case, in the form of a causal relative risk. Our ensemble machine learning is optimized for prediction, but it does not directly provide measures of individual variable importance. We conducted follow-up procedure (TMLE) to generate interpretable estimates of variable impact with robust standard errors [32, 33].

## Results

Descriptive results show the age distribution of laboratory-confirmed patients across the three different epidemic phases **(Supplemental Figure S2**). Phases 1 and 2 have similar distributions, and there are more young people (under 30) in phase 3. The six most prevalent pre-existing conditions are hypertension, obesity, diabetes, smoking, asthma, and renal disease **(Supplemental Figure S3**). The prevalence of all pre-existing conditions decreased over the three phases, and prevalence of hypertension, obesity, and diabetes were drastically reduced in phase 3.

### Super learner (SL) prediction

SL fit has high prediction accuracy on the testing set (AUC: 0.907 (95% CI: (0.905-0.908)). SL leverages XGBoost models (**Supplemental Table S2)** and significantly outperforms the simple logistic regression model (testing AUC: 0.874 (95% CI: (0.872-0.876)) (**Table 2**). The logistic regression model overpredicts mortality risks for those roughly above age 75 compared to the SL prediction (**Fig. 1**). Permuted variable importance shows, while holding other variables constant, age is consistently the most important for SL prediction in average mortality risk (**Supplemental Figure S4 and Table S3)**. Having multiple comorbidities can dramatically increase risk for those individuals (**Fig. 2**).

**Table 2.**
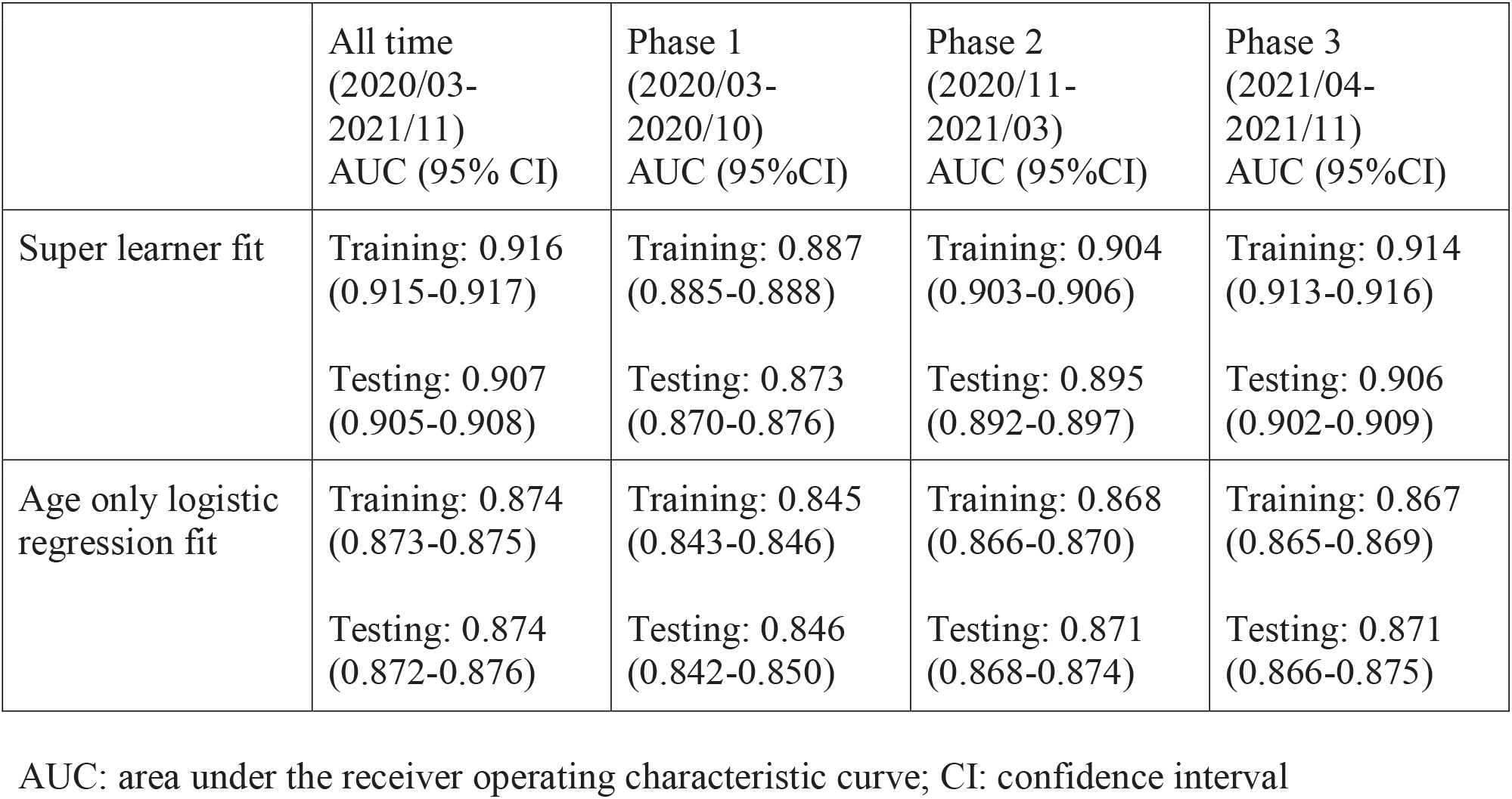
Prediction results.

**Fig. 1.**
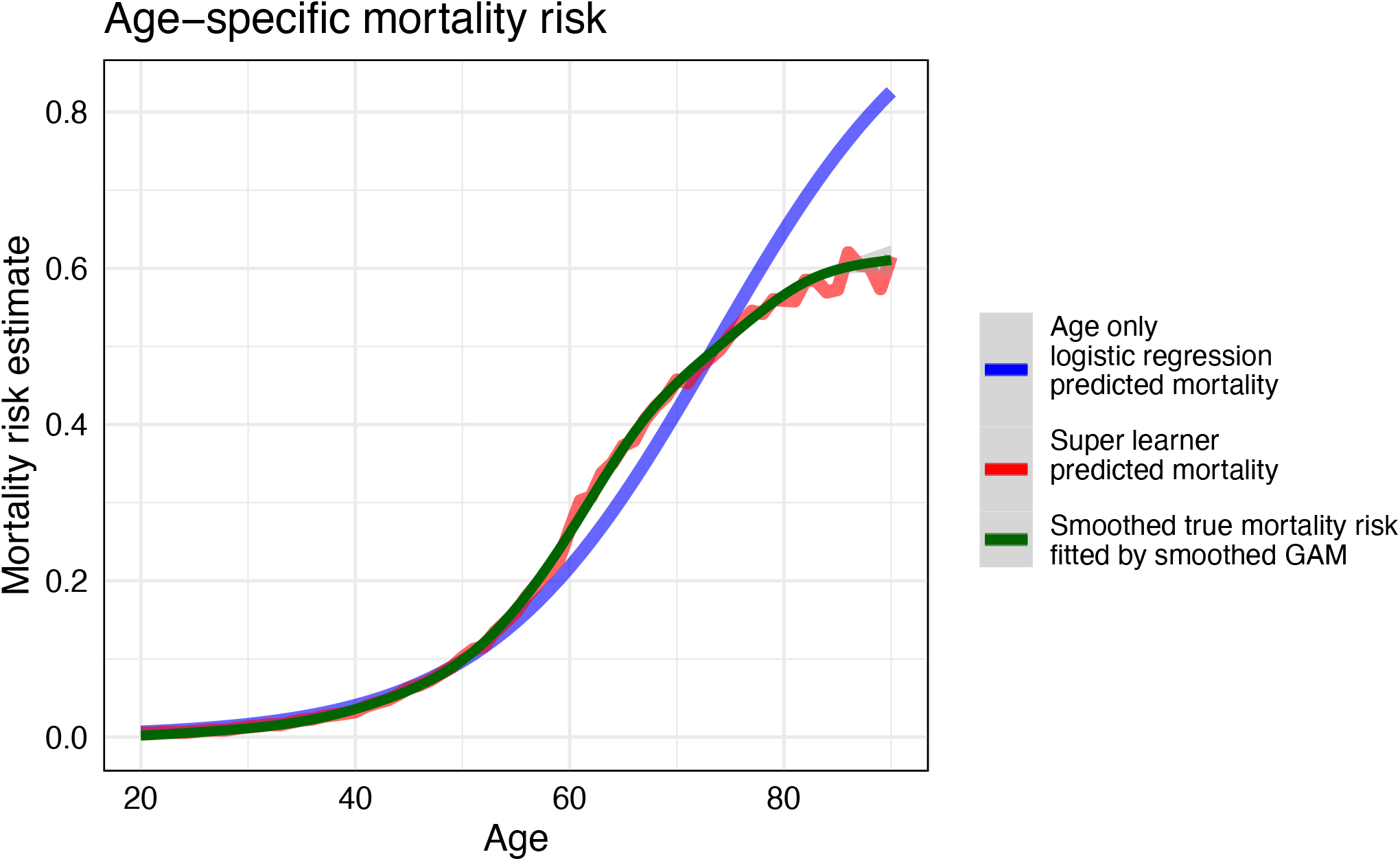
Mortality risk prediction comparing age only logistic regression and super learner. GAM: generalized additive model The smoothed true mortality risk curve is generated using a GAM with integrated smoothness estimation fitted with cubic splines.

**Fig. 2.**
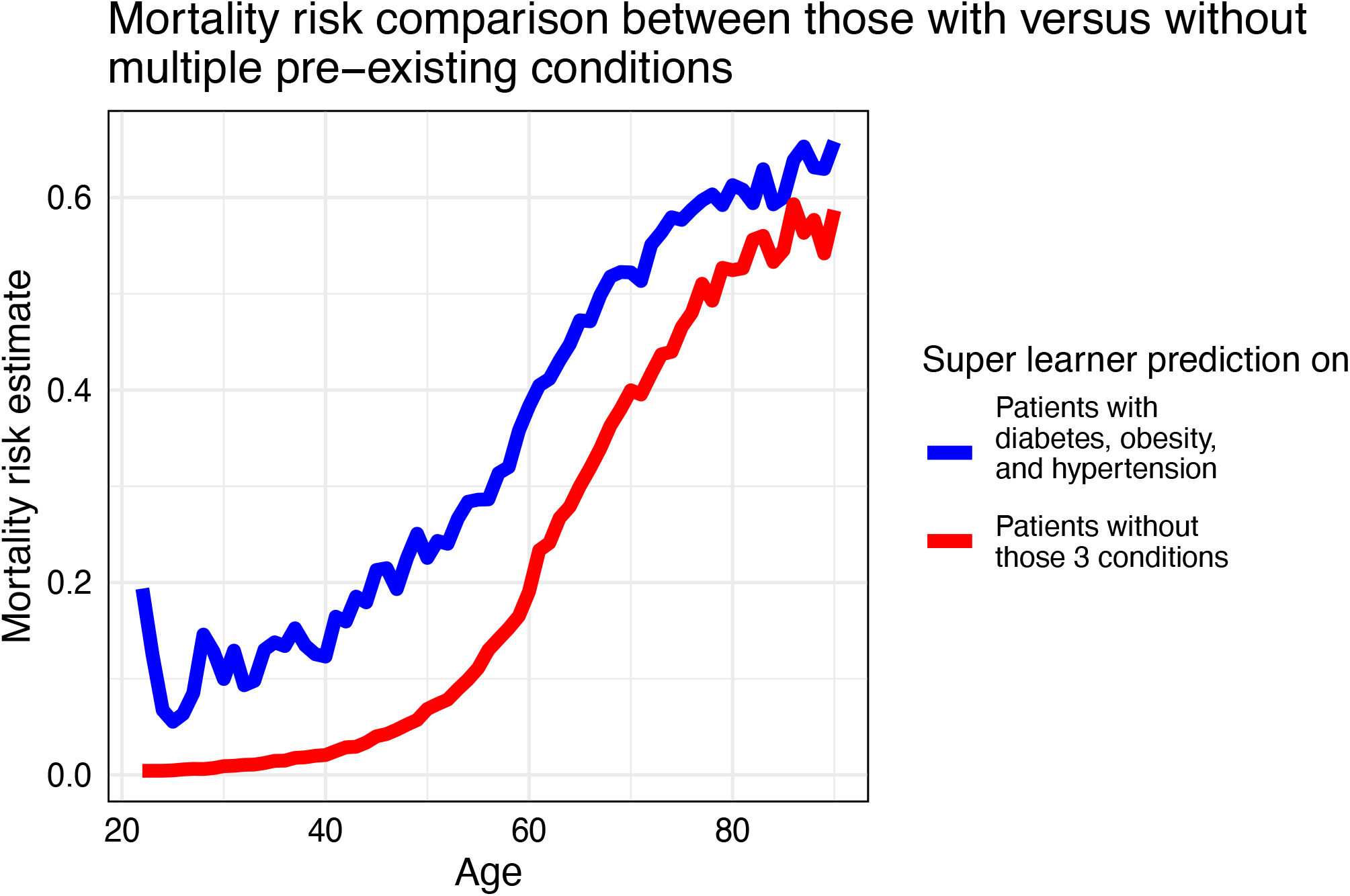
Super learner predicted mortality risk averaged by specific age in two subgroups: those having all obesity, diabetes, and hypertension pre-existing conditions versus those without.

### Relative risks of pre-existing conditions

To assess the impact of each pre-existing condition, we estimate their respective relative risks (RRs) of mortality, adjusting for demographic variables. We report the estimated RRs in **Table 3**, ordered by impact (most to least) (**Supplemental Figure S5**). The RRs compare the expected risk if all patients have the pre-existing condition (with) versus if all patients do not have the condition (without). The highest impact pre-existing condition is renal disease (RR: 3.783, 95% CI: (3.705, 3.862)); diabetes, obesity, and hypertension also have high impact individually (RR: 1.432-1.847). Minimal differences between the risk estimates are shown for smoking and asthma (RR: 1.049 and 1.037, respectively).

**Table 3.** Targeted maximum likelihood estimation relative risk results for each pre-existing condition.

|  | All time<br>(2020/03-2021/11)<br>Relative Risk<br>(95% CI) | Phase 1<br>(2020/03-2020/10)<br>Relative Risk<br>(95%CI) | Phase 2<br>(2020/11-2021/03)<br>Relative Risk<br>(95%CI) | Phase 3<br>(2021/04-2021/11)<br>Relative Risk<br>(95%CI) |
| --- | --- | --- | --- | --- |
| Renal disease | 3.783<br>(3.705, 3.862) | 2.588<br>(2.521, 2.657) | 2.994<br>(2.910, 3.080) | 6.638<br>(6.361, 6.927) |
| Diabetes | 1.847<br>(1.820, 1.875) | 1.536<br>(1.508, 1.566) | 1.594<br>(1.564, 1.625) | 2.508<br>(2.423, 2.596) |
| Hypertension | 1.745<br>(1.721, 1.770) | 1.427<br>(1.402, 1.452) | 1.500<br>(1.474, 1.527) | 2.356<br>(2.279, 2.436) |
| Obesity | 1.432<br>(1.417, 1.447) | 1.269<br>(1.249, 1.288) | 1.259<br>(1.239, 1.279) | 1.794<br>(1.750, 1.840) |
| Smoking | 1.049<br>(1.030, 1.068) | 1.001<br>(0.975, 1.028) | 0.992<br>(0.966, 1.018) | 1.158<br>(1.107, 1.210) |
| Asthma | 1.037<br>(1.002, 1.073) | 0.941<br>(0.895, 0.989) | 0.942<br>(0.892, 0.995) | 1.223<br>(1.134, 1.319) |

CI: confidence interval

The phase analyses indicate pre-existing conditions are especially important in phase 3. Phase 1 and 2 are very similar in terms of both risk prediction and adjusted mortality risk estimates. However, in phase 3, age is less important in prediction (**Supplemental Table S3)** and RRs drastically increase for every comorbidity. The adjusted risks show the decrease for each pre-existing condition in phase 3 (**Supplemental Table S4)**.

## Discussion

Our analysis of (>1.4 million) laboratory-confirmed COVID-19 patients demonstrates that age is by far the most important predictor of average mortality. For those patients with renal disease, diabetes, hypertension, or obesity, having the comorbidity further increases their risk of mortality. A patient with diabetes, hypertension, and obesity is roughly comparable to a patient 20 years older with none of the conditions, based on the predicted mortality (**Fig. 2**). Thus, having a comorbidity increases risk of mortality and should be considered at any age. The reason that comorbidities add little to the predictive power at younger ages is that hypertension and diabetes are age-related and the reported onset is often for those over 30, so the pre-existing conditions are far less prevalent.

Our prediction results using machine learning methods predict better than previous studies, and we demonstrated the feasibility and robustness of using machine learning methods targeted for prediction and variable impact. SL model prediction has an AUC of 0.907, which is higher than any previous Mexican study (AUCs from 0.634 to 0.824) [8,34]. Although age has been well reported by previous studies as important [5,34,35], our analysis is more robust because we do not assume a pre-specified functional relationship between the explanatory variables and the predicted variable, and thereby avoid any arbitrary groupings into age categories. Moreover, since those above age 60 have a higher prevalence of comorbidities, relying on simple logistic regression models can greatly overpredict the average mortality risk for the elder patients. Our study applies TMLE to estimate the adjusted mortality risk ratios for each comorbidity to provide more robust impact estimates that respect time ordering and account for background variables.

We find consistent results of comorbidities compared to previous studies, and present phase analyses highlighting the changes in relative risks over time. Previous results from logistic regressions indicated odds ratios of 1.458-2.48 for renal disease, 1.237-1.74 for diabetes, 1.173-1.47 for obesity, 1.194-1.315 for hypertension, 0.852-1.02 for smoking, and 0.74-1.420 for asthma [34–36]. Although our analysis is generally consistent with previous findings, our RR estimations have less uncertainty. Renal disease has the greatest impact on mortality, followed by diabetes, hypertension, and obesity; smoking and asthma have negligible impact on mortality risk.

This phase-specific analysis produced a seemingly paradoxical finding. The impact of comorbidities on predicted mortality decreased with time (primarily between the second and third wave), but the RR on mortality dramatically increased for the same conditions **(Supplemental Table S4 and Figure S5)**. The apparent explanation is that mortality risk for people without the comorbidities fell faster than for people with them, increasing the relative risk. The decrease in mortality risk is multifactorial and includes a decrease in susceptibility over time (due to prior infection and vaccination), improved treatment, enhanced healthcare response and opportunity to be admitted to a hospital or ICU, and less virulent viral subtypes. This implies that as herd immunity increases, medical resources should focus even more on protecting vulnerable people at older age and those with comorbidities since they are even more likely to experience severe outcomes compared to those who are younger and/or healthier.

Readers should be cautious about extrapolating our findings to other populations. Although our sample is large and includes patients from all parts of Mexico, most of the patients were IMSS beneficiaries. In order to access IMSS health services, patients require: a) be a formal-sector worker or retired, b) be a direct dependent of such an employee, c) be a bachelor or postgraduate student in a public institution, d) voluntarily enroll by paying a fee. Thus, the IMSS population skews toward the upper half of the income distribution. Populations without similar access to health services may have different results. It is also important to consider the potential impact of data quality. Pre-existing conditions were self-reported and likely also inconsistently recorded, perhaps in systematic ways that could have biased the results. For example, if people with severe diabetes were more likely to report diabetes as a pre-existing condition, we may overestimate the impact of diabetes on mortality.

It is also important to consider what predictive variables are included in this model. We sought to predict risk for an individual in the population using their characteristics prior to infection. In other words, what is this person’s risk of death from COVID-19 if they were to be infected? The answer to this question best informs the question of who should be prioritized for protection against infection or for early therapeutic interventions following infection. It does not attempt to predict the likely mortality of a patient who presents to the health services with COVID-19 because information about that patient’s severity of their COVID-19-related symptoms will represent important additional predictors of their mortality risk.

## Data Availability

The study was conducted using confidential patient records subject to strict access controls and we are therefore unable to share the data that were used for this study.

## Abbreviations

AUC: area under the receiver operating characteristic curve
CI: confidence interval
COPD: chronic obstructive pulmonary disease
COVID-19: coronavirus disease of 2019
IMSS: Mexican Social Security Institute
PCR: polymerase chain reaction
RR: relative risk
SL: super learner
TMLE: targeted maximum likelihood estimation
XGBoost: extreme gradient boosting

## Acknowledgements

We thank the staff of C3.ai DTI for their technical support and our colleagues at University of California, Berkeley, the Mexican National Autonomous University, and the Mexican Social Security Institute (IMSS) for all of the administrative and technical support that has allowed this collaboration to flourish.

## Declarations

### Authors’ contributions

LDL and AEH contributed to the study design and methodology. AJ, YY, and IA contributed to data acquisition. LDL, YY, and KK contributed to data cleaning. LDL led the data analysis and visualization. LDL, AEH, JPG, and SMB interpreted the results. LDL drafted the manuscript with support from RG on literature search. AEH and SMB significantly contributed to the revision of the manuscript. All authors participated in review and edited the manuscript; all authors have read and approved the final manuscript. All authors had full access to all the data in the study and accepted responsibility to submit for publication. All authors take responsibility for the integrity of the data and the accuracy of the data analysis.

### Ethics approval and consent to participate

This data-only study was approved on November 4th, 2020, by the Scientific Research National Committee (Social Security Mexican Institute) with R-2020-785-165. The University of California, Berkeley Institutional Review Board (IRB) determined that the project was exempt from IRB approval.

### Consent for publication

Not applicable.

## Supplemental Material

<u>SupplementalMaterial.pdf</u>:

Table S1. TableS1 – [Complete table of baseline variables and pre-existing conditions]

Table S2. TableS2 – [Weighted combination of the super learner fit]

Table S3. TableS3 – [Top 5 ranked most important variables for prediction]

Table S4. TableS4 – [Targeted maximum likelihood estimation adjusted mortality risk, with or without the pre-existing condition]

Figure S1. FigS1 – [Flowchart for analytic sample development]

Figure S2. FigS2 – [Age distribution for laboratory-confirmed COVID-19 patients]

Figure S3. FigS3 – [Prevalence of pre-existing conditions prevalence over time]

Figure S4. FigS4 – [Prediction variable importance predicted using the super learner fit]

Figure S5. FigS5 – [Relative risk for each pre-existing condition associated with mortality]

