## Supplemental for "Who is most at risk of dying if infected with SARS-CoV-2? A mortality risk factor analysis using machine learning of COVID-19 patients over time in a large Mexican population"

**Table S1. Complete table of baseline variables and pre-existing conditions**

| | All time (2020/03-2021/11) | Phase 1 (2020/03-2020/10) | Phase 2 (2020/11-2021/03) | Phase 3 (2021/04-2021/11) |
|---|---|---|---|---|
| sample size | 1,423,720 | 303,278 | 425,698 | 694,744 |
| Demographic variables | | | | |
| Age in years (mean (SD)) | 42.15 (15.70) | 46.41 (16.04) | 44.89 (16.27) | 38.61 (14.34) |
| Sex = male (%) | 729,782 (51.3) | 158,248 (52.2) | 218,165 (51.2) | 353,369 (50.9) |
| Insured by IMSS = 1(%) | 1,358,440 (95.4) | 288,588 (95.2) | 402,754 (94.6) | 667,098 (96.0) |
| Indigenous = 1 (%) | 7,381 (0.5) | 2,200 (0.7) | 1,628 (0.4) | 3,553 (0.5) |
| Year-month patient initiated care | | | | |
| 2020/03 | 1,061 (0.1) | 1,061 (0.3) | 0 (0.0) | 0 (0.0) |
| 2020/04 | 10,832 (0.8) | 10,832 (3.6) | 0 (0.0) | 0 (0.0) |
| 2020/05 | 30,720 (2.2) | 30,720 (10.1) | 0 (0.0) | 0 (0.0) |
| 2020/06 | 51,079 (3.6) | 51,079 (16.8) | 0 (0.0) | 0 (0.0) |
| 2020/07 | 60,780 (4.3) | 60,780 (20.0) | 0 (0.0) | 0 (0.0) |
| 2020/08 | 49,618 (3.5) | 49,618 (16.4) | 0 (0.0) | 0 (0.0) |
| 2020/09 | 44,758 (3.1) | 44,758 (14.8) | 0 (0.0) | 0 (0.0) |
| 2020/10 | 54,430 (3.8) | 54,430 (17.9) | 0 (0.0) | 0 (0.0) |
| 2020/11 | 65,437 (4.6) | 0 (0.0) | 65,437 (15.4) | 0 (0.0) |
| 2020/12 | 93,748 (6.6) | 0 (0.0) | 93,748 (22.0) | 0 (0.0) |
| 2021/01 | 145,858 (10.2) | 0 (0.0) | 145,858 (34.3) | 0 (0.0) |
| 2021/02 | 68,421 (4.8) | 0 (0.0) | 68,421 (16.1) | 0 (0.0) |
| 2021/03 | 52,234 (3.7) | 0 (0.0) | 52,234 (12.3) | 0 (0.0) |
| 2021/04 | 35,181 (2.5) | 0 (0.0) | 0 (0.0) | 35,181 (5.1) |

| | | | | |
|---|---|---|---|---|
| 2021/05 | 26,300 (1.8) | 0 (0.0) | 0 (0.0) | 26,300 (3.8) |
| 2021/06 | 45,986 (3.2) | 0 (0.0) | 0 (0.0) | 45,986 (6.6) |
| 2021/07 | 170,212 (12.0) | 0 (0.0) | 0 (0.0) | 170,212 (24.5) |
| 2021/08 | 249,477 (17.5) | 0 (0.0) | 0 (0.0) | 249,477 (35.9) |
| 2021/09 | 116,569 (8.2) | 0 (0.0) | 0 (0.0) | 116,569 (16.8) |
| 2021/10 | 48,515 (3.4) | 0 (0.0) | 0 (0.0) | 48,515 (7.0) |
| 2021/11 | 2,504 (0.2) | 0 (0.0) | 0 (0.0) | 2,504 (0.4) |
| Mexican states (%) | | | | |
| Aguascalientes | 26,420 (1.9) | 6,897 (2.3) | 12,350 (2.9) | 7,173 (1.0) |
| Baja California | 43,925 (3.1) | 13,677 (4.5) | 14,188 (3.3) | 16,060 (2.3) |
| Baja California Sur | 24,521 (1.7) | 4,300 (1.4) | 5,423 (1.3) | 14,798 (2.1) |
| Campeche | 9,557 (0.7) | 1,728 (0.6) | 765 (0.2) | 7,064 (1.0) |
| CDMX 1 Noroeste | 32,552 (2.3) | 5,374 (1.8) | 13,174 (3.1) | 14,004 (2.0) |
| CDMX 2 Noreste | 54,249 (3.8) | 11,370 (3.7) | 20,273 (4.8) | 22,606 (3.3) |
| CDMX 3 Suroeste | 42,896 (3.0) | 9,701 (3.2) | 17,588 (4.1) | 15,607 (2.2) |
| CDMX 4 Sureste | 62,097 (4.4) | 13,248 (4.4) | 25,899 (6.1) | 22,950 (3.3) |
| Chiapas | 14,826 (1.0) | 2,836 (0.9) | 1,801 (0.4) | 10,189 (1.5) |
| Chihuahua | 23,229 (1.6) | 6,489 (2.1) | 6,879 (1.6) | 9,861 (1.4) |
| Coahuila | 48,933 (3.4) | 16,355 (5.4) | 15,459 (3.6) | 17,119 (2.5) |
| Colima | 18,310 (1.3) | 2,997 (1.0) | 2,633 (0.6) | 12,680 (1.8) |
| Durango | 19,738 (1.4) | 6,228 (2.1) | 6,674 (1.6) | 6,836 (1.0) |
| Guanajuato | 61,570 (4.3) | 11,595 (3.8) | 29,274 (6.9) | 20,701 (3.0) |
| Guerrero | 23,871 (1.7) | 4,502 (1.5) | 3,920 (0.9) | 15,449 (2.2) |
| Hidalgo | 23,673 (1.7) | 4,658 (1.5) | 8,266 (1.9) | 10,749 (1.5) |
| Jalisco | 104,054 (7.3) | 19,491 (6.4) | 27,868 (6.5) | 56,695 (8.2) |

| | | | | |
|---|---|---|---|---|
| Mexico Oriente | 108,067 (7.6) | 20,368 (6.7) | 39,633 (9.3) | 48,066 (6.9) |
| Mexico Poniente | 48,973 (3.4) | 11,632 (3.8) | 17,026 (4.0) | 20,315 (2.9) |
| Michoacan | 30,570 (2.1) | 6,246 (2.1) | 7,221 (1.7) | 17,103 (2.5) |
| Morelos | 18,797 (1.3) | 2,845 (0.9) | 6,817 (1.6) | 9,135 (1.3) |
| Nayarit | 23,934 (1.7) | 3,378 (1.1) | 2,994 (0.7) | 17,562 (2.5) |
| Nuevo Leon | 105,912 (7.4) | 23,776 (7.8) | 30,114 (7.1) | 52,022 (7.5) |
| Oaxaca | 24,324 (1.7) | 4,493 (1.5) | 5,391 (1.3) | 14,440 (2.1) |
| Puebla | 50,998 (3.6) | 9,287 (3.1) | 17,932 (4.2) | 23,779 (3.4) |
| Queretaro | 41,977 (2.9) | 4,707 (1.6) | 19,259 (4.5) | 18,011 (2.6) |
| Quintana Roo | 38,390 (2.7) | 4,607 (1.5) | 4,542 (1.1) | 29,241 (4.2) |
| San Luis Potosi | 26,118 (1.8) | 7,353 (2.4) | 7,573 (1.8) | 11,192 (1.6) |
| Sinaloa | 44,333 (3.1) | 11,030 (3.6) | 9,405 (2.2) | 23,898 (3.4) |
| Sonora | 27,691 (1.9) | 7,245 (2.4) | 6,083 (1.4) | 14,363 (2.1) |
| Tabasco | 16,004 (1.1) | 2,622 (0.9) | 1,719 (0.4) | 11,663 (1.7) |
| Tamaulipas | 38,941 (2.7) | 8,504 (2.8) | 7,442 (1.7) | 22,995 (3.3) |
| Tlaxcala | 13,809 (1.0) | 2,769 (0.9) | 4,957 (1.2) | 6,083 (0.9) |
| Veracruz Norte | 41,804 (2.9) | 10,002 (3.3) | 6,801 (1.6) | 25,001 (3.6) |
| Veracruz Sur | 35,019 (2.5) | 10,292 (3.4) | 4,970 (1.2) | 19,757 (2.8) |
| Yucatan | 35,126 (2.5) | 5,968 (2.0) | 5,091 (1.2) | 24,067 (3.5) |
| Zacatecas | 18,512 (1.3) | 4,708 (1.6) | 8,294 (1.9) | 5,510 (0.8) |
| Pre-existing conditions | | | | |
| Asthma = yes (%) | 25,297 (1.8) | 7,951 (2.6) | 7,765 (1.8) | 9,581 (1.4) |
| Cardiovascular disease = yes (%) | 17,816 (1.3) | 6,643 (2.2) | 6,389 (1.5) | 4,784 (0.7) |
| Chronic liver disease = yes (%) | 1,875 (0.1) | 710 (0.2) | 668 (0.2) | 497 (0.1) |
| COPD = yes (%) | 15,390 (1.1) | 5,825 (1.9) | 5,496 (1.3) | 4,069 (0.6) |

| | | | | |
|---|---|---|---|---|
| Diabetes = yes (%) | 169,869 (11.9) | 55,551 (18.3) | 61,120 (14.4) | 53,198 (7.7) |
| Hemolytic anemia = yes (%) | 705 (0.0) | 276 (0.1) | 246 (0.1) | 183 (0.0) |
| HIV = yes (%) | 4,717 (0.3) | 1,133 (0.4) | 1,425 (0.3) | 2,159 (0.3) |
| Hypertension = yes (%) | 228,901 (16.1) | 72,615 (23.9) | 83,735 (19.7) | 72,551 (10.4) |
| Immunosuppression = yes (%) | 10,434 (0.7) | 4,102 (1.4) | 3,453 (0.8) | 2,879 (0.4) |
| Neurological disease = yes (%) | 1,645 (0.1) | 544 (0.2) | 559 (0.1) | 542 (0.1) |
| Obesity = yes (%) | 181,736 (12.8) | 55,965 (18.5) | 60,217 (14.1) | 65,554 (9.4) |
| Smoking = yes (%) | 87,161 (6.1) | 21,253 (7.0) | 28,346 (6.7) | 37,562 (5.4) |
| Cancer diagnosis = yes (%) | 3,751 (0.3) | 1,178 (0.4) | 1,317 (0.3) | 1,256 (0.2) |
| Renal disease diagnosis = yes (%) | 24,099 (1.7) | 8,912 (2.9) | 8,555 (2.0) | 6,632 (1.0) |
| Tuberculosis = yes (%) | 675 (0.0) | 203 (0.1) | 218 (0.1) | 254 (0.0) |

CDMX: Ciudad de México; COPD: Chronic obstructive pulmonary disease; HIV: human immunodeficiency virus; SD: standard deviation
Mexican states refer to where the patient was treated

**Table S2. Weighted combination of the super learner fit**

| Machine learning candidate algorithm | Weights | Mean squared error | Standard error |
|---|---|---|---|
| Bayesian additive regression trees | 0 | 0.266 | 0.0002 |
| Bayesian generalized linear model | 0 | 0.067 | 0.0003 |
| Elastic net regression | 0 | 0.068 | 0.0004 |
| Empirical mean | 0 | 0.094 | 0.0005 |
| XGBoost (multiple tuning) | 0.596 (combined) | 0.065 (on average) | 0.0003 (on average) |
| Generalized additive model | 0.222 | 0.066 | 0.0004 |
| LASSO regression | 0 | 0.067 | 0.0004 |
| Logistic regression | 0 | 0.067 | 0.0004 |
| Multivariate Adaptive Regression Splines | 0 | 0.068 | 0.0004 |
| Random forest | 0.181 | 0.066 | 0.0002 |
| Ridge regression | 0 | 0.067 | 0.0002 |

LASSO: least absolute shrinkage and selection operator; XGBoost: extreme gradient boosting
XGBoost coefficients are combined; mean squared error and standard error were averaged.

**Table S3. Top 5 ranked most important variables for prediction**

| | All time (2020/03-2021/11) | Phase 1 (2020/03-2020/10) | Phase 2 (2020/11-2021/03) | Phase 3 (2021/04-2021/11) |
|---|---|---|---|---|
| Rank 1 | Age 0.147 | Age 0.209 | Age 0.208 | Age 0.069 |
| Rank 2 | Year-month patient initiated care 0.014 | Renal disease 0.008 | Mexican state 0.007 | Renal disease 0.004 |
| Rank 3 | Renal disease 0.005 | Sex 0.007 | Renal disease 0.006 | Mexican state 0.003 |
| Rank 4 | Mexican state 0.004 | Year-month patient initiated care 0.007 | Insured by IMSS 0.006 | Diabetes 0.003 |
| Rank 5 | Sex 0.004 | Mexican state 0.006 | Sex 0.006 | Insured by IMSS 0.002 |

IMSS: Mexican Institute of Social Security
Mexican state refers to where the patient was treated.
Measured by the log-likelihood difference in prediction pre-post permutation of each variable while holding all others constant.

**Table S4. Targeted maximum likelihood estimation adjusted mortality risk, with or without the pre-existing condition**

| | All time (2020/03-2021/11) | | Phase 1 (2020/03-2020/10) | | Phase 2 (2020/11-2021/03) | | Phase 3 (2021/04-2021/11) | |
|---|---|---|---|---|---|---|---|---|
| | with | without | with | without | with | without | with | without |
| Renal disease | 0.381 | 0.101 | 0.439 | 0.170 | 0.425 | 0.142 | 0.305 | 0.046 |
| Diabetes | 0.173 | 0.094 | 0.247 | 0.161 | 0.214 | 0.135 | 0.104 | 0.041 |
| Hypertension | 0.162 | 0.093 | 0.231 | 0.162 | 0.201 | 0.134 | 0.097 | 0.041 |
| Obesity | 0.141 | 0.099 | 0.212 | 0.168 | 0.177 | 0.141 | 0.080 | 0.045 |
| Smoking | 0.110 | 0.105 | 0.176 | 0.176 | 0.146 | 0.147 | 0.056 | 0.048 |
| Asthma | 0.109 | 0.105 | 0.166 | 0.177 | 0.139 | 0.147 | 0.059 | 0.049 |

**Fig. S1. Flowchart for analytic sample development**

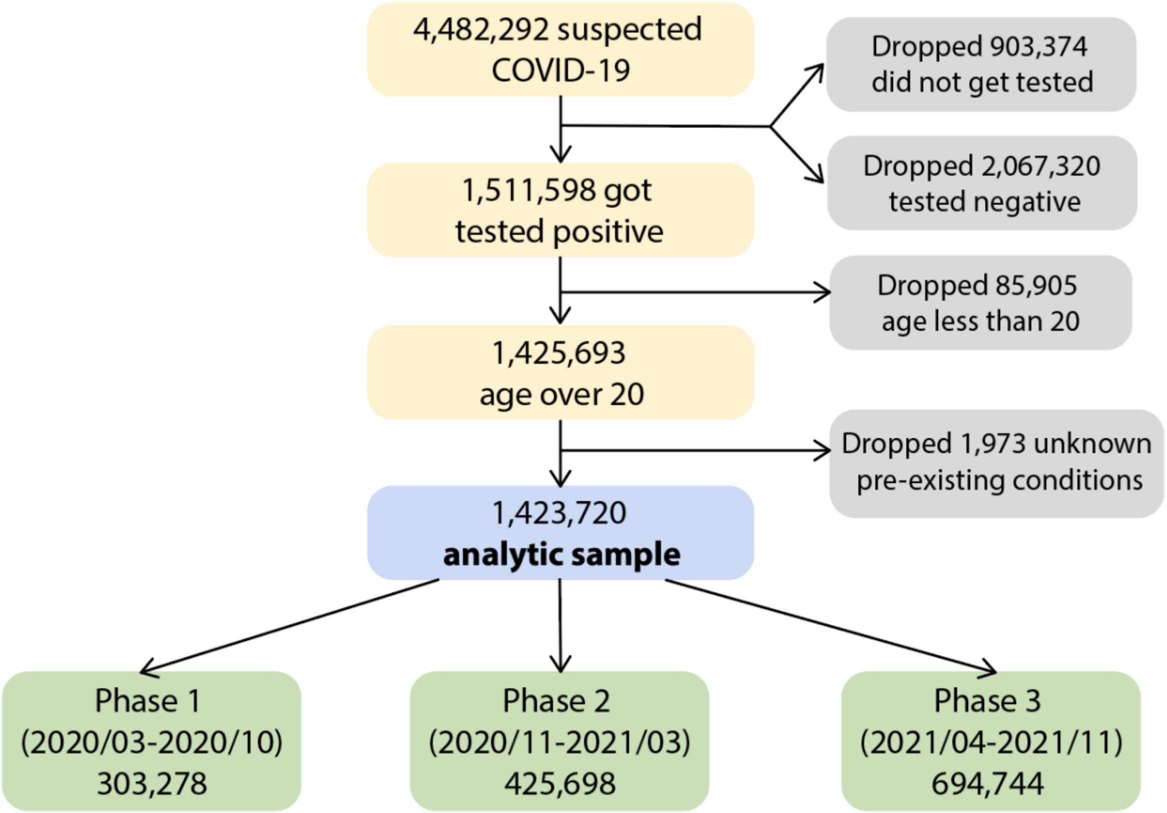

**Fig. S2. Age distribution for laboratory-confirmed COVID-19 patients**

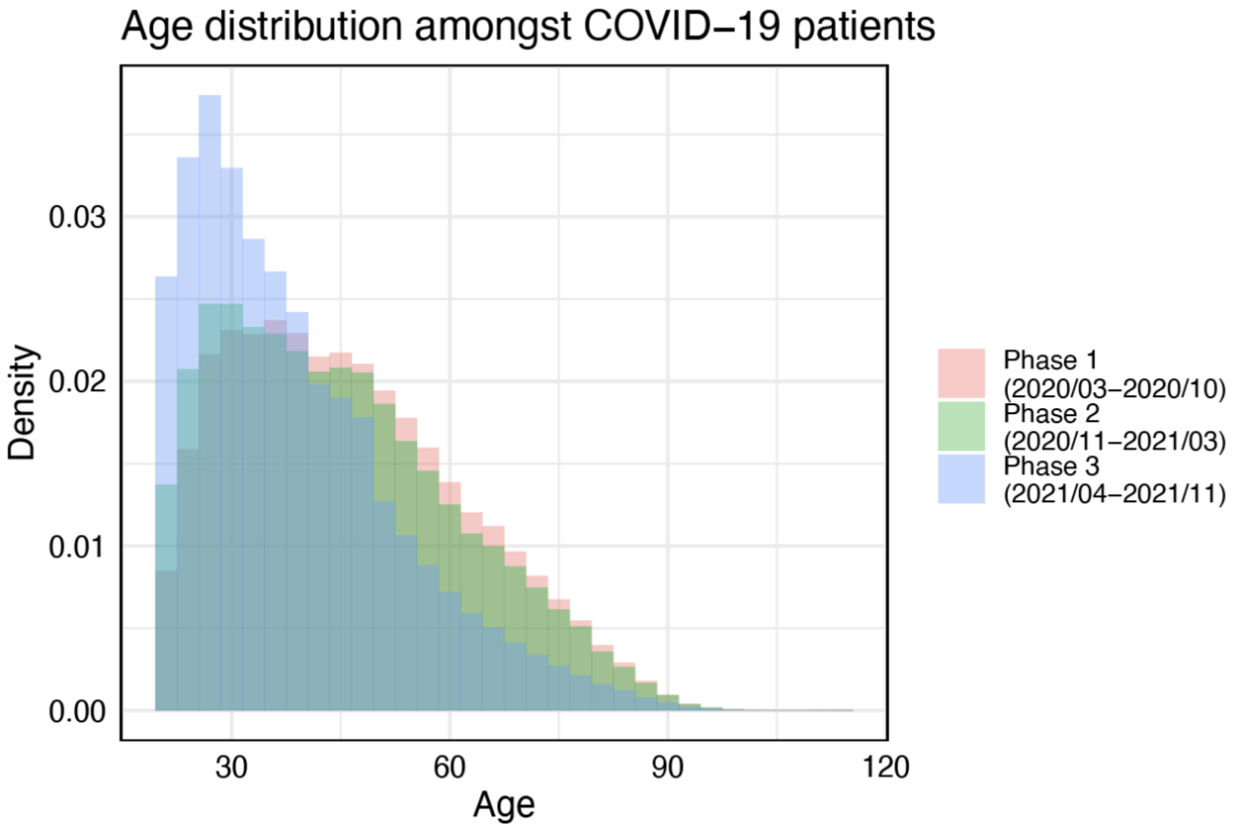

**Fig. S3. Prevalence of pre-existing conditions prevalence over time**

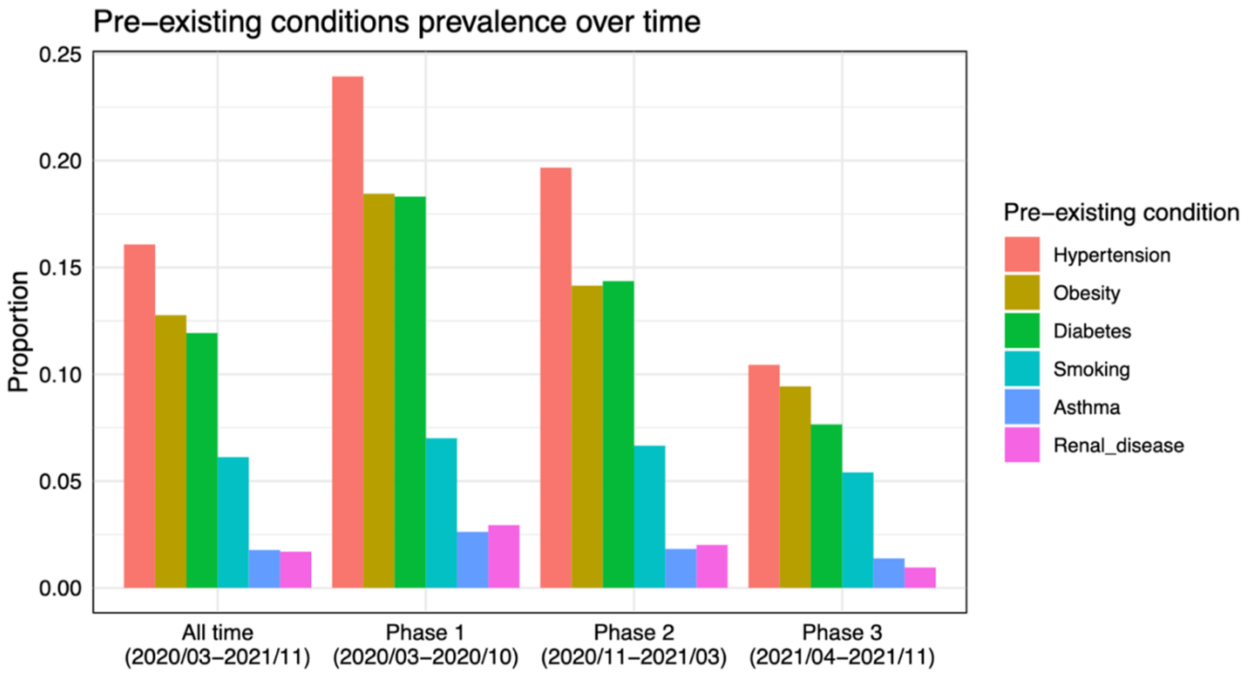

**Fig. S4. Prediction variable importance predicted using the super learner fit**

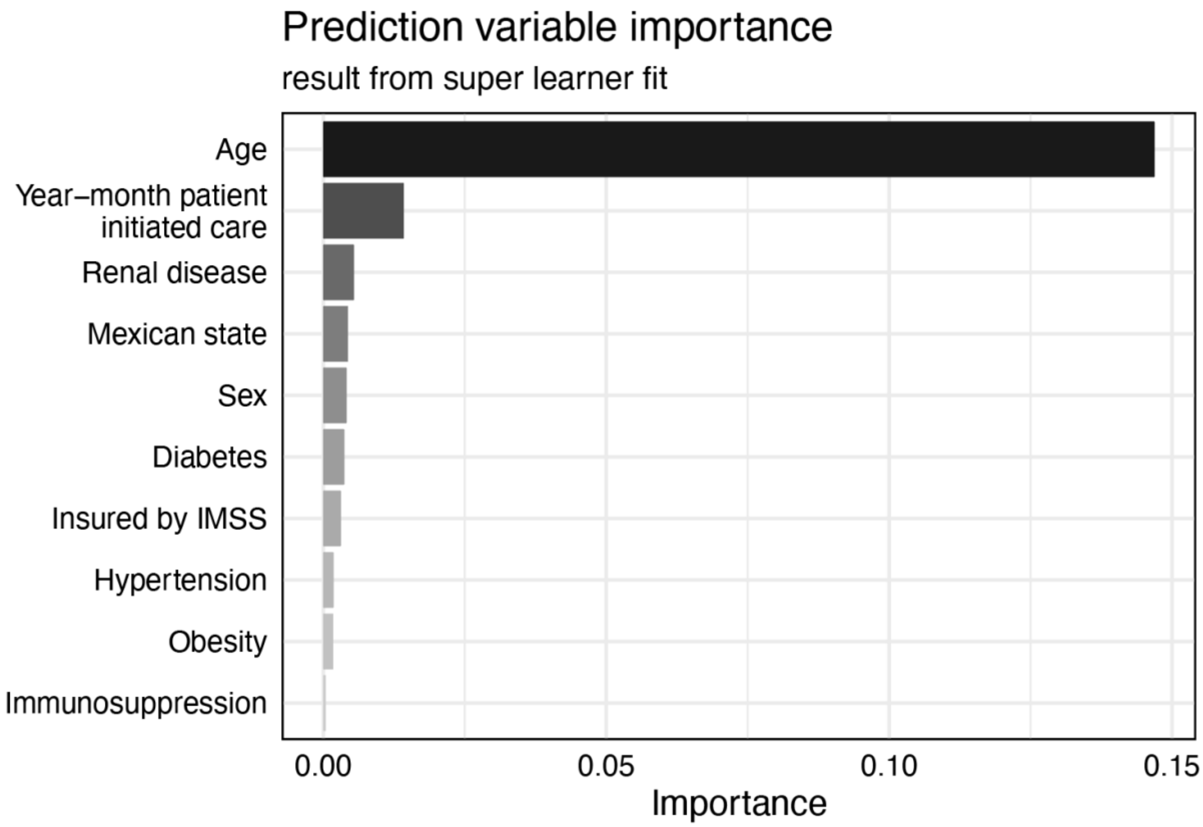

IMSS: Mexican Institute of Social Security

**Fig. S5. Relative risk for each pre-existing condition associated with mortality**

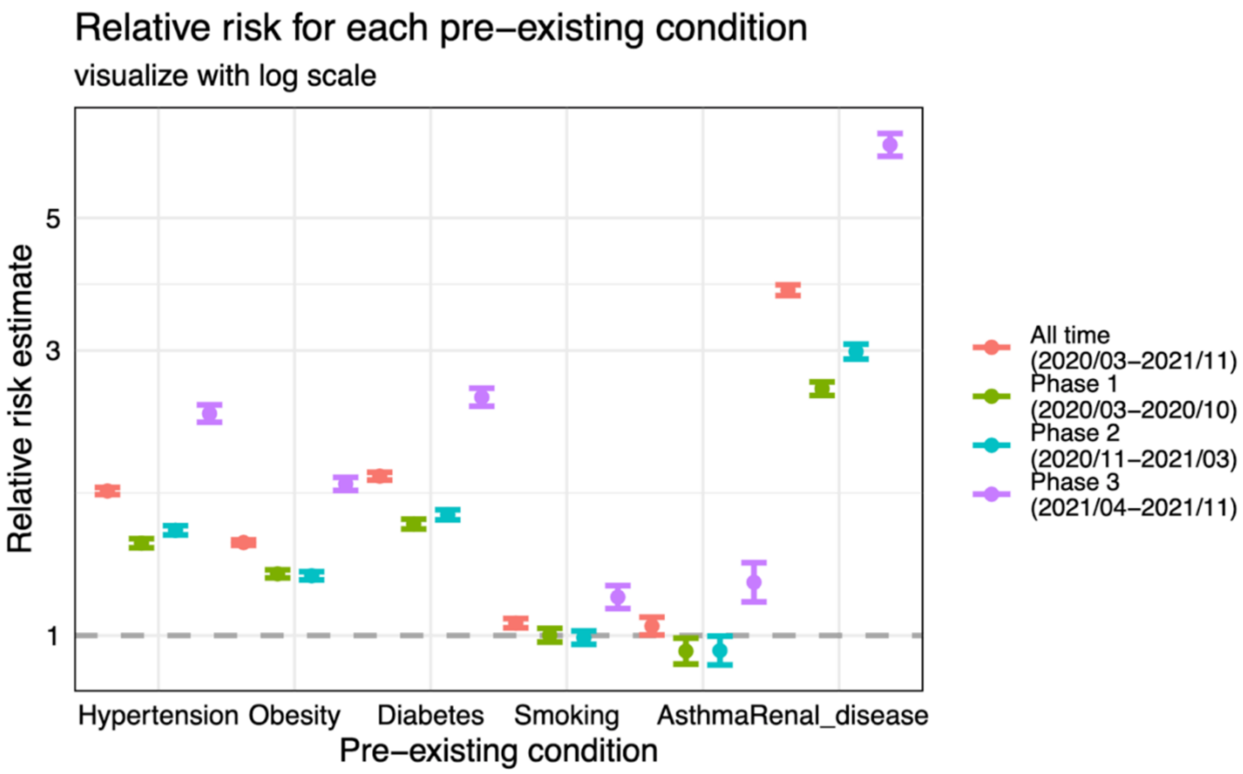

Error bars reflect a 95% confidence interval.
